# Neurocognition After Perturbed Sleep (NAPS): A Protocol for an Experimental Study Examining the Impact of Sleep on Neurocognition, Functioning, and Clinical Indicators in People with Schizophrenia

**DOI:** 10.1101/2025.08.18.25333917

**Authors:** SA Miller, LH Ospina, AE Mullins, A Parekh, K Kam, CR Fowler, AW Varga, D Kimhy

**Affiliations:** Department of Psychiatry, Icahn School of Medicine at Mount Sinai, New York, NY, USA; Mount Sinai Integrative Sleep Center, Division of Pulmonary, Critical Care, and Sleep Medicine, Icahn School of Medicine at Mount Sinai, New York, NY, USA; Department of Mathematical Sciences, Worcester Polytechnic Institute, Worcester, MA, USA

**Author notes:** Please address all correspondence to: Dr. David Kimhy Department of Psychiatry Icahn School of Medicine at Mount Sinai One Gustave L. Levy Place, Box 1230 New York, NY 10029.

## Abstract

**Introduction:** People with schizophrenia (SZ) display neurocognitive deficits that have been identified as predictors of poor functioning and disability. Previous reports have noted that these deficits are strikingly similar to the cognitive sequalae of poor sleep. Consistent with this observation, extensive animal and basic human research have strongly support the deleterious effects of poor sleep on neurocognition. Notably, insomnia and disturbed sleep (DS) are highly ubiquitous among people with SZ. Yet, there is scarce information about the potential impact of poor sleep on neurocognition in this population, with available studies using correlational or cross-sectional designs.

**Method:** We will employ an experimental, within-person, cross-over design to characterize sleep architecture, duration, continuity and quality along with their neurocognitive, functional, electrophysiological, and clinical sequalae in individuals with SZ. Participants will complete two overnight lab-based polysomnography examinations (two weeks apart) employing two sleep schedules: 1) undisturbed sleep (8 hours); and 2) perturbed sleep (4 hours). As part of each overnight sleep schedule, participants will complete brief EEG-indexed sleep-sensitive memory tasks pre- and post-sleep, a post-sleep neurocognitive test battery, as well as a three-day post-sleep digital phenotyping of physical activity, daily functioning, mood, symptoms, medication side-effects, and suicidal ideation via smartphones and actigraphy.

**Discussion:** Despite their chronic and ubiquitous nature, the impact of insomnia and DS on neurocognition in SZ remain poorly understood and modeled. This lack of information hinders our ability to develop accurate treatment models and effective interventions for neurocognitive deficits in SZ. Our aim is to address these gaps in knowledge.

## Background

People with schizophrenia (SZ) display a range of neurocognitive deficits across multiple domains including processing speed, attention, verbal and visual memory, executive function, working memory, and social cognition^1–4^. These deficits have been identified as predictors of poor functioning and disability^2,6–8^, thus representing a serious public health concern and an important target for interventions^9,10^. Previous reports have noted that the neurocognitive deficits observed in people with SZ are strikingly similar to the cognitive sequalae of poor sleep^11^. Consistent with this observation, extensive evidence from animal^12–14^ and basic human research^15–17^ strongly support the deleterious effects of poor sleep on neurocognition. In humans, evidence points to poor sleep resulting in deterioration in multiple neurocognitive domains including cognitive control^18–21^, attention^22–27^, constructive thinking^16^, working memory^28–31^, as well as verbal memory^32^, a meta-analysis of 70 studies (n=1458) pointing to moderate-to-large effect sizes for attention and working memory^33^. Relatedly, these deficits parallel findings of poor sleep resulting in altered synaptic properties in the prefrontal cortex^34,35^ and in the hippocampus^34,36–38^, including hippocampal neurogenesis suppression^39,40^.

Extensive basic research has implicated Slow Wave Sleep (SWS) as a critical process associated with neurocognition. Often referred to as “deep sleep”, SWS consists of Stage 3 of non-rapid eye movement (NREM) sleep, characterized by low frequency, high amplitude oscillatory EEG SWS activity^41^. Evidence from rodent studies indicate hippocampal activity during exploration was subsequently replayed during SWS^42,43^, suggesting that SWS replay strengthens neural connections underlying learning. In humans, imaging studies indicates that SWS-related blood flow to the right hippocampus is associated with spatial navigation task improvement^44^, and augmentation using transcranial current oscillating (at a frequency mimicking SWS during sleep) enhanced declarative memory retention^45^. Another sleep process relevant to neurocognition is spindle activity, characterized as amplitude bursts of neural oscillatory activity (at frequency between 11-15 Hz) that are generated by the interplay of the thalamic reticular nucleus and other thalamic nuclei during Stage 2 NREM sleep^46–48^. Findings suggest spindles facilitate synaptic plasticity and memory consolidation^41,47,49–52^, specifically declarative memory^53^, the integration of new information into existing knowledge^54^, and directed remembering and forgetting^55^.

Germane to SZ, insomnia and disturbed sleep (DS) are highly ubiquitous in this population, with prevalence reaching up to 80%, a rate far higher than in the general population^56,57^. Specifically, an extensive research literature employing polysomnographic, actigraphy, and self-report measures point to increased sleep latency, greater wake time after sleep onset, along with reduced total sleep time and consolidation^56,58,59^ in drug-naïve and medicated individuals with SZ^60^, as well as individuals at clinical high risk for psychosis^61^. Several reviews have highlighted the negative influence of sleep disruption on neurocognition in SZ^62–64^, pointing to associations between SWS and impaired neurocognitive performance in medication-naïve^65^, medicated^66–69^, and unmedicated individuals with SZ^70^. A number of neurocognitive domains have been found to be negatively impacted including attention^65,71^, procedural learning^69,72^, visuospatial memory^68^, and declarative memory^67,68^. These findings have been also documented among individuals at risk for psychosis^73^. In domains other than memory, longer SWS was found to be correlated with quicker problem solving^74^ and enhanced attention^75^. Likewise, reduction of sleep spindle activity has been linked with impaired neurocognition in individuals with SZ^76–79^. Specifically, a recent review suggested reduced sleep spindle activity is associated with impaired consolidation of memory^77^, as well as impairments in attention^65,80^ and verbal cognition^81^. Notably, a small treatment study of Eszopiclone, a sedative-hypnotic medication used to treat insomnia, increased sleep spindle density in SZ, but not sleep-dependent memory^82^. Additionally, poor neurocognition in SZ has been linked to alterations in a number of biomarkers including inflammation markers^83–85^ and Brain-Derived Neurotrophic Factor (BDNF)^86,87^. Our group has shown cognitive deficits were associated with higher peripheral TNF-α and Interleukin 12p70^83^.

Numerous studies have shown sleep deprivation also results in alterations in neurocognition-related biomarkers, specifically inflammatory markers^88^ including increases in TNF-α^89^, IL-1β, IL-1 receptor antagonist, as well as decreases in CRP and IL-6^90^. Sleep deprivation also decreases BDNF secretion, which plays a central role in neuroplasticity and neurocognition^27,91–93^, including in SZ^86^. Clinical reports also point to sleep difficulties having been implicated in deleterious impact on emotion regulation and stress,^94–96^ increased risk for mood disorders^97–101^, as well as contributing to exacerbation of psychotic symptoms in both clinical and non-clinical populations,^61,63,102,103^ ^104–110^ including hallucinations^111–116^ and paranoia^117–119^.

Converging lines of animal, preclinical and clinical research suggest that insomnia and DS may play a significant role in neurocognitive functioning among people with SZ. However, the research literature has a number of critical limitations: 1) the extant studies have employed cross-sectional and/or correlational designs; 2) a plurality of sleep and neurocognition in SZ studies have employed actigraphy and/or retrospective self-reports of sleep, limiting the ability to ascertain underlying neurobiological mechanisms; 3) previous studies typically centered on sleep’s impact on a single or narrow range of neurocognitive domains (i.e., memory); 4) there are virtually no data quantifying the impact of sleep on daily functioning in SZ; 5) sleep has been linked to neurocognition-related biomarkers (e.g., BDNF, inflammation markers), however there is scarce data on their impact in SZ; 6) finally, there have been no experimental studies examining the impact of insomnia and DS on neurocognition in SZ, limiting the ability to determine whether they are a cause or an effect of clinical phenomena. Thus, despite their chronic and ubiquitous nature, insomnia and DS remain poorly understood and modeled in SZ, their impact is rarely considered in clinical trials, and they remain largely unaddressed by clinicians. This lack of information hinders our ability to develop accurate treatment models and effective interventions for neurocognitive deficits in SZ.

To address this gap in the literature, the primary aim of this study is to conduct a comprehensive characterization on the putative links between sleep, neurocognition, functioning, and other clinical indicators in individuals with SZ. We will characterize insomnia and DS in individuals with SZ and quantify their impact on neurocognition and functioning. Employing an experimental, within-person, repeated assessment design, we will characterize sleep architecture, duration, continuity and quality along with their neurocognitive, electrophysiological, biomarkers and functional sequalae in individuals with SZ. Participants will first complete clinical measures and a week-long, at-home characterization of sleep duration and quality using actigraphy and a sleep diary. Next, they will complete two overnight lab-based polysomnography examinations (two weeks apart) employing two sleep schedules: 1) undisturbed sleep (8 hours); and 2) perturbed sleep (4 hours). As part of these schedules, participants will complete brief EEG-indexed memory tasks pre- and post-sleep, along with a post-sleep neurocognitive test battery. Finally, following each overnight sleep, participants will complete three consecutive days of digital phenotyping of mood, emotion regulation, symptoms, medication side-effects, social functioning, and suicidal ideation via smartphones.

## Methods

### Design

The inclusion criteria were age 18-60 years; a DSM-5 diagnosis of SZ, schizoaffective, or schizophreniform disorder; taking antipsychotic medication for ≥8 weeks and on current doses for 4 weeks, and/or injectable depot antipsychotics with no change in the last 3 months; and capacity to understand all the potential risks and benefits of the study. The exclusion criteria were a DSM-5 alcohol/substance disorder diagnosis (except nicotine) within the last 6 months; taking medications affecting sleep propensity or architecture (other than psychotropic medication); initiation of medications known to impact cognition in previous 4 weeks or any change in doses during this period; history of seizures/head trauma with loss of consciousness (>10 min) resulting in cognitive sequelae; medical or neurological conditions that could interfere with study participation (e.g., mental retardation, narcolepsy, REM behavior disorder, parasomnias); pregnant/nursing; serious homicidal/suicidal risk (past 6 months); “moderate” or more severe disorganization (PANSS≥4); poor English reading ability (WTAR<7); being employed as vehicle drivers/train operators or have occupations in which lapses in sustained vigilance would compromise safety; being employed as a night shift worker or having an irregular sleep-wake rhythms (based on the week-long home actigraphy; i.e., average bedtime of 11pm±2 hours); participation in a neurocognition study in the past 3 months.

**Figure 1.**
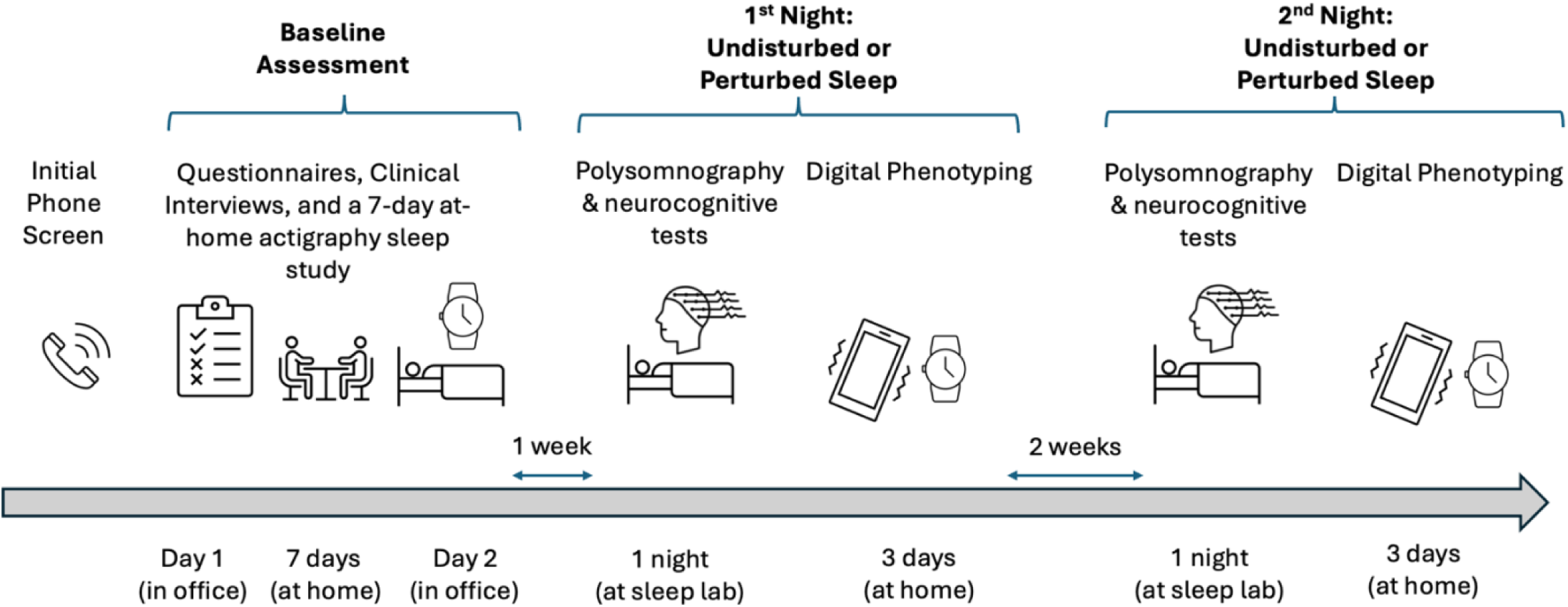
Timeline of Research Assessments **Note:** Participants were randomized to either undisturbed (ad-lib) sleep or perturbed sleep condition for their 1^st^ in-lab polysomnography night and completed the other condition during the 2^nd^ night.

Following an initial phone screen, participants completed a baseline assessment over two visits. On Day 1, participants completed interview-based assessments of diagnoses and symptoms (SCID-5 and PANSS), a reading ability test (WTAR), as well as assessment of substance use (via urine toxicology), smoking, and pregnancy. At the end of Day 1, participants were provided with an Actigraph wGT3X-BT monitor to wear for a week-long characterization of at-home sleep. Participants returned the actigraph on Day 2, scheduled a week after Day 1, and completed measures of sleep history, daytime sleepiness, and emotion regulation.

Individuals meeting the full inclusion/exclusion criteria were then invited to complete two overnight, physician-supervised polysomnography assessments at a sleep lab. We employed two sleep schedules: 1) undisturbed sleep (8 hours); and 2) perturbed sleep (4 hours). The choice of 4-hour sleep time reflected a balance between a period of sleep that would likely negatively impact neurocognitive functioning and other clinical indicators, while also considering participants’ safety. The 4-hour sleep period may also represent a relatively common “real-world” experience of many individuals, contributing the generalizability of the findings. The order of the sleep schedules was determined by random assignment, with the assessments scheduled 2 weeks apart. All polysomnography assessments were completed on Mondays/Tuesdays to allow participants to complete three weekdays of digital phenotyping following each polysomnography assessment.

For each polysomnography assessment, participants arrived at the sleep lab at 7:30pm, at which time they were acclimatized to their private bedroom. The windowless rooms included a bed, a private bathroom with a shower, a TV and access to Wi-Fi. Participants were allowed to bring and use personal electronic devices (e.g., smartphones, iPads). Following the acclimation period, participants were connected to the polysomnography/EEG equipment (∼60 min) and completed pre-sleep EEG-indexed sleep-sensitive neurocognitive tasks (∼60 min). From ∼9:30pm participants were allowed to engage in unstructured activities in their room (e.g., reading, watching TV) until sleep time. Participants randomized to the undisturbed sleep were instructed to go to sleep at 11pm. Participants randomized to the perturbed sleep were instructed to remain awake until 3am. A technician monitored all participants during their stay and ensured sleep/wake status.

In the morning, participants in both sleep schedules were awoken by the lab technician at 7am and ate a standardized small breakfast (coffee/tea and a power bar). At 7:30am, participants repeated post-sleep EEG-indexed sleep-sensitive neurocognitive tasks (∼60 min). Next, at ∼8:30am, after being disconnected from the polysomnography/EEG equipment, participants completed a neurocognitive battery (MCCB; ∼60 min). Finally, at 10am participants were released home to complete a 3-day “real-world” digital phenotyping assessment of mood, emotion regulation, symptoms, side-effects, daily functioning, physical activity, and suicidal ideation via smartphones employing Experience Sampling Method (ESM) and Actigraph monitor wGT3X-BT.

Participants’ sleep undisturbed/perturbed schedules were randomized using an a-priori randomization schedule list. All clinical raters had a master’s degree in psychology or higher and completed rater training in administration and scoring of all cognitive, clinical, and functioning assessments (*i.e.*, MCCB, SCID, PANSS) prior to study initiation. Participants were compensated up to US$420 for completing all research assessments.

Typically, participants completed the research protocol over 5-6 weeks. All participants continued to receive their regular psychiatric and medical care, and no effort was made to adjust/influence treatments.

### Measures

Table 1 presents the assessment domains, measures, administration time, and study phase of administration. The measures include:

1. Demographics and Reading Ability: Data collected included race, gender, ethnicity, age, education, employment, language fluency, marital status, and current living arrangements. Reading ability was indexed by the Wechsler Reading Ability Test (WTAR).
2. Diagnoses: were established using a modified version of the Structured Clinical Interview for the DSM, 5^th^ edition (SCID-V).
3. Sleep Characteristics:
a. Home Sleep Schedule and Characteristics: Participants were provided with an Actigraph monitor wGT3X-BT to wear for a week-long, in-home characterization of habitual “real-world” sleep/wake times to determine exclusion criterion, including total sleep time, sleep efficiency and wake after sleep onset. Sleep history and daytime sleepiness was assessed using the Epworth Sleepiness Scale (ESS)^120^; and Pittsburgh Sleep Quality Index (PSQI)^121^.
b. Polysomnography: EEG sleep studies were used to characterize sleep latency, duration, continuity, and architecture overnight. Respiratory airflow was measured from a nasal cannula. Polysomnograms were scored in 30-second epochs using standard criteria (American Academy of Sleep Medicine; AASM)^122,123^ for sleep and EEG arousals. Total sleep time and percent time spent in wake, non-REM stage 1 (NREM 1), non-REM stage 2 (NREM 2), non-REM stage 3 (NREM 3), and REM sleep were determined. Respiratory events were scored from the airflow signal using AASM criteria and stage specific (REM, NREM 1, NREM 2, NREM 3, and Total) apnea indices. Apneas and hypopneas (with both oxygen desaturation and arousal) were scored according to AASM standards and apnea-hypopnea indices (AHI) were calculated. Continuity of sleep was assessed as duration of sleep runs (consecutive 30-second epochs of sleep scored at that stage, terminated by an epoch scored as another stage). To determine SWS activity, EEG data from the experimental nights were imported into MATLAB and epoched into 30 sec bins. Epochs containing artifacts were rejected, and the remaining epochs were filtered between 0.4 and 50 Hz. A fast Fourier transform was then applied to the filtered EEG signal at 30 sec intervals with 50% overlap employing a Hamming window. Absolute SWS activity in an EEG lead was defined as the spectral power between 0.5 and 4.0 Hz during non-REM sleep. Relative SWS activity was defined as the absolute

**Table 1.** Assessment Domains, Measures, Administration Time, and Phase of Administration.

| Domain | Measure | Admin Time | Screen/<br>Baseline | 1<sup>st</sup><br>Night | 2<sup>nd</sup><br>Night |
| --- | --- | --- | --- | --- | --- |
| Phone Screen | Phone Screen form | 5 min | + |  |  |
| Demographics | Demographics form | 5 min | + |  |  |
| Reading Ability | Wechsler Reading Ability Test (WTAR) | 5 min | + |  |  |
| Diagnoses | Structured Clin Interview for the DSM, 5<sup>th</sup> ed. (SCID-5) | 60 min | + |  |  |
| Symptoms | Positive & Negative Syndrome Scale (PANSS) | 45 min | + |  |  |
|  | Beck depression Inventory (BDI-II) | 5 min | + |  |  |
|  | Beck Anxiety Inventory | 5 min | + |  |  |
| Suicide Risk | The Beck Scale for Suicide Risk (BSS) | 5 min | + |  |  |
| Substance Use,<br>Smoking and<br>Pregnancy | Urine Toxicology Test & Pregnancy Test | 5 min | + |  |  |
|  | Fagerström Test for Nicotine Dependence (FTND) | 5 min | + |  |  |
| Medications | Medications Form | 5 in | + |  |  |
| Emotion<br>Awareness &<br>Regulation | Toronto Alexithymia Scale (TAS-20) | 5 min | + |  |  |
|  | Emotion Regulation Questionnaire (ERQ) | 5 min | + |  |  |
| Sleep | Epworth Sleepiness Scale (ESS) | 5 min | + |  |  |
|  | Pittsburgh Sleep Quality Index (PSQI) | 5 min | + |  |  |
|  | Actigraphy | -- | + * | + *** | + *** |
|  | Polysomnography | -- |  | + ** | + ** |
| Neurocognition | MATRICS Consensus Cognitive Battery (MCCB) | 65 min |  | + | + |
|  | Virtual Maze Navigation Task | 20 min |  | + | + |
|  | Word Memory Task | 20 min |  | + | + |
|  | Emotional Memory Task | 20 min |  | + | + |
| Digital<br>Phenotyping | “Real World” Daily Functioning | -- |  | + *** | + *** |

Note: \* 7-day at-home assessment; \*\* 1-night sleep lab assessment; \*\*\* 3-day ambulatory assessments via actigraphy and smartphones employing Experience Sampling Method indexing physical activity, daily functioning, mood, emotion regulation, symptoms, medication side effects, and suicidal risk.

SWS activity divided by the absolute spectral power across all frequencies (0.5-50 Hz) for EEG lead, allowing spectral power normalization across participants.

4. Neurocognition:
a. Overall neurocognitive functioning was indexed by the composite score of the MATRICS Consensus Cognitive Battery (MCCB)^1,2^. The MCCB also provides domain scores indexing speed of processing, attention/vigilance, working memory, verbal learning, visual learning, reasoning, problem solving, and social cognition.
b. Additionally, we employed three tasks sensitive to the impact of disturbed sleep:
i. The Virtual Maze Navigation Task: a computer-generated 3D virtual maze designed to index overnight consolidation of spatial navigational memory, with sensitivity to both reduced SWS and REM sleep^124–128^ . The game was projected onto a screen in a darkened testing room. The viewing area was 67”x 50”, and participants sat 13’ away with vision corrected (if required). In the pre-sleep administration, after a period of general familiarization with joystick controls (ThrustmasterTM) in a Z-shaped corridor, participants spent 5 min exploring a maze using “Unreal Tournament 3 Editor”^124^. Avatar walking and turning speed was reduced to minimize motion “cyber-sickness.” Participants were instructed to remember the layout of the maze environment. Next, they navigated through the same maze during three pre-sleep test trials in which they were instructed to reach a specified goal point as quickly as possible. Time to reach the goal per trial was capped at 600 sec. In the post-sleep administration, participants first performed a standardized 10-min psychomotor vigilance test (PVT) employing a vigilance task monitor^129^ to control for impact of sleep on attention. For the PVT, participants held an electronic device on which a counter is programmed to begin at random intervals. Participants were instructed to press a button as quickly as they are aware the counter has begun^124–126,129–133^. Performance metrics on the psychomotor vigilance test included reaction time (in milliseconds) and number of lapses (no response after 500 millisec). As the number of lapses typically has a skewed distribution, transformation of the number of lapses was applied to enable parametric testing (lapses transform=sqrt(#lapses)+sqrt (#lapses+1))^129^. Following the PVT, participants performed three final test trials on the 3D virtual maze, again instructed to reach the same specified goal point as quickly as possible. Performance metrics on the virtual maze included completion time (CT), distance traveled within the maze, and distance backtracking. Change was calculated as the difference between the averages across the 3 trials pre- and post-sleep, normalized to the average of the 3 trials pre-sleep for each metric. Participants were counterbalanced for the maze encountered first in a pseudo-random fashion.
ii. Word Memory Task: The episodic word-pair task, adapted from Mander et al., 2013 (based on Diekelmann & Born, 2010; Marshall et al., 2006)^131–133^ involves encoding and recognition of word-pairs pre- and post-sleep recollection. It was selected based on its sensitivity to reduced SWS activity^131^. The task used word-nonsense word pairs to maximize the novel episodic and associated hippocampal-dependent demands of the task^134^ and minimize the semantic, and therefore hippocampal-independent, demands of the task^134–136^. Participants initially completed pre-sleep intentional encoding phase, followed immediately by a criterion phase. Next, following a brief delay, participants completed a short recognition test (10 min; 30 studied trials and 15 foil trials). Following overnight sleep, participants completed a long delay recognition test (90 studied trials and 45 foil trials). Words were 3–8 letters in length and drawn from a normative set of English words^131^. Nonsense words were 6–14 letters in length, derived from groups of common phonemes^134^. The word-pair task began pre-sleep with an encoding phase composed of 120 word-nonsense word trials.

During each encoding trial, a word-nonsense word pair was shown for 5 sec., with criterion training occurring immediately after encoding. During each self-paced criterion trial, a previously studied probe word was presented with its original nonsense word associate from encoding and two new nonsense words not previously shown. After responding, the participants were given feedback for 1 sec, with incorrect responses resulting in trial repetition at random intervals. During recognition trials post-sleep, either a previously studied probe word or a new (foil) probe word was shown for 5 sec with four response options presented below. When a previously studied probe word was presented, the following response options were available: 1) the nonsense word originally paired with that probe word at encoding (hit); 2) a previously studied nonsense word, but one presented with a different previously studied probe word (lure); 3) a new nonsense word never seen during encoding; and 4) an option designating the shown probe word as new. New nonsense words were only presented once during the entire experiment, whereas previously studied nonsense words were presented twice during recognition, always in the same session, as a lure on one trial and the correct paired associate on another. When a foil probe word was presented, the four response options consisted of three new nonsense words never presented during learning (which, if chosen, would be a false alarm), and an option designating this foil probe word as new (which would be a correct rejection). Associative recognition memory was calculated by subtracting the false alarm rate (% of foil words endorsed as previously studied) and the lure rate (% of previously studied words recognized but erroneously paired with the lure and hence incorrect nonsense word associate) from the hit rate (% of previously studied probe words accurately paired with the correct nonsense word associate)^134^. Episodic memory change was calculated as the difference in short- and long-delay recognition memory scores^131^.

iii. Emotional Memory Task (Hammerer Emotional Memory Task^137^): The stimuli consisted of 120 indoor and outdoor pictures containing negative emotional or neutral scenes partly taken from the International Affective Picture System (IAPS) database^138^ (N_negative_=48, N_neutral_=27) and partly taken from an image set collected from the Internet (N_negative_=72, N_neutral_=93). The Internet-based image set was built as part of a different study and was rated on valence and arousal by a sample of 60 young adults (mean age=28±2 years, 50% female). The Self-Assessment Manikin^139^ was used, which is an affective rating system devised by Lang (1980) that also underlies IAPS ratings. For each image to be rated, participants could select from a 9-point rating scale with 9 representing a high rating on each dimension (i.e., high pleasure, high arousal) and 1 representing a low rating on each dimension (i.e., low pleasure, low arousal). The rating was performed on a total of 387 novel images collected from the Internet as well as 45 IAPS images. For the current study, we chose negative emotional pictures with low valence and moderately high arousal, along with neutral pictures with neutral valence and low arousal. Stimuli were displayed on a 22-inch monitor and viewed from a distance of 80 cm. The stimuli, fixation crosses, and the gray-patterned background were adjusted for luminance to control for trivial luminance-related effects on PD responses. Stimuli and fixation crosses were displayed in the center of the screen. The text was presented in size 50 Arial font and colored white. Participants sat on a comfortable chair, with their head position stabilized with a desktop-mounted chin and headrest. Two 4-choice button boxes were used to record responses during the task.

Taking advantage of the infrastructure of the study, we also collected the following clinical indicators:

1. Symptoms: were indexed by the Positive & Negative Syndrome Scale (PANSS)^140^ and the Beck Inventories (BDI-II/BAI)^141,142^. The PANSS is a 30-item semi-structured interview assessing positive symptoms, negative symptoms, and general psychopathology. The BDI-II is a 21-item self-report measure indexing depression. The BAI is a 21-item self-report questionnaire indexing anxiety.
2. Digital Phenotyping: Following each overnight assessment, participants completed 3-day digital phenotyping via smartphones employing Experience Sampling Method (ESM) to explore the impact of sleep on “real-world” mood, emotion regulation, symptoms, medication side effects, daily functioning, and suicidal ideation. Our group has published the first study validating the use of digital phenotyping with electronic mobile devices in SZ^143^. The ESM questions were adapted from another NIMH funded psychosis study (“*Using mHealth To Optimize Pharmacotherapy Regimens*”, a sub-Project of “*Optimizing and Personalizing Interventions for Schizophrenia Across the Lifespan*”; P50MH115843; PIs: Stroup & Kimhy).
3. Emotion Awareness and Regulation: The Toronto Alexithymia Scale (TAS-20)^144,145^ and Emotion Regulation Questionnaire (ERQ)^146,147^ were used to quantify emotion awareness and regulation, respectively. The TAS-20 is a 20-item self-report questionnaire, with higher scores indicating poorer functioning. We used the Difficulty Identifying Feelings (DIF; 7 items) and Difficulty Describing Feelings (DDF; 5 items) subscales. Participants are asked to indicate on a 5-point scale to what extent they agreed with each statement. The ERQ is a 10-item self-report questionnaire assessing reappraisal (6 items) and suppression (4 items). Participants are asked to indicate on a seven-point scale (from 1=“strongly disagree” to 7=“strongly agree”) to what extent they agree with each statement, with higher scores reflecting stronger endorsement of using the strategy. Previous reports by our group have found emotion awareness and regulation to be strong predictors of social functioning and psychotic symptoms in people with schizophrenia and at risk for psychosis^148–151^.
4. Suicide Risk: The Beck Scale for Suicide Risk (BSS)^152^ is a self-report 19-item scale preceded by five screening items. The BSS and its screening items are intended to assess a patient’s thoughts, plans and intent to commit suicide.
5. Substance Use and Smoking: Substance use was determined via urine toxicology test. Smoking was indexed by the Fagerström Test for Nicotine Dependence (FTND)^153^.
6. Medications: were recorded based on self-report. Antipsychotic medications were indexed by chlorpromazine equivalence. Other classes of medications recorded include: 1) antidepressants; 2) mood stabilizers; and 3) other medications known to impact sleep and or cognition (e.g., PRN antihistamines).

## Discussion

Converging lines of research support the premise that poor sleep may have a detrimental effect on neurocognition and functional outcomes in individuals diagnosed with SZ. However, the extant sleep and neurocognition in SZ research literature has been limited by several critical methodological barriers. The present protocol aims to address these very limitations via the use of an experimental design to probe the links between sleep and neurocognition employing state-of-the-art measures to assess multiple domains of neurocognition and functioning. If confirmed, the results of this study may have significant implications for future treatment of neurocognition in patients with SZ by highlighting sleep as a key target for the development of novel interventions to treat neurocognitive deficits in individuals with SZ.

## Data Availability

All data produced are available online at NIMH Data Archive (ID#: C3934)

https://nda.nih.gov

## Protocol Registration and Archiving

The protocol was registered in ClinicalTrials.gov (ID #NCT05032963). The neurocognitive and related data from the protocol were archived at the NIMH Data Archive (ID #3934).

